# Association between types of physical activity and risk of ischemic heart disease based on US Guidelines: A systematic review and meta-analysis

**DOI:** 10.1101/2023.03.06.23286885

**Authors:** Jaewoo Cha, Jeehyun Kim, Kwan Hong

**Affiliations:** Department of Preventive Medicine, Korea University College of Medicine, Seoul, Republic of Korea

**Author notes:** **Corresponding author:** Jaewoo Cha, MPH, Department of Preventive Medicine, Korea University College of Medicine, 73, Goryeodae-ro, Seongbuk-gu, Seoul 02841, Republic of Korea.

## Abstract

**Background:** Mortality from ischemic heart disease (IHD) is increasing worldwide. There is no available literature regarding the validity of US guidelines for IHD risk reduction through physical exercise. In this meta-analysis, we aimed to measure the effectiveness of US guidelines for physical activity in reducing IHD risk based on the types of exercise.

**Methods:** Six databases, including MEDLINE, EMBASE, Cochrane Library, CINAHL, Scopus, and Web of Science, were searched from January 1, 2000, to November 6, 2022. The most recent literature search was conducted on November 6, 2022, and only English-language articles were included. Studies designed and conducted on humans on any type of IHD-related physical activity were included. Study outcomes included heterogeneity among the studies, overall effects of all types of physical activity, and IHD risk. The random effects model was measured. A funnel plot was used to assess publication bias.

**Results:** When the US guidelines for physical activity were not applied, there was a high level of heterogeneity in the effects of physical activity by type, with overall effects of 0.764 (odds ratio [OR], 95% confidence interval [CI]: 0.737–0.795). The effect of physical activity on IHD was 0.593 (OR, 95% CI: 0.489–0.720). No publication bias was observed. After applying the US guidelines for physical activity, there was a low level of heterogeneity in the effects of physical activity by type and an observed OR of 0.515 (95% CI: 0.401–0.662) for myocardial infarction.

**Discussion:** Each type of exercise had different effects on reducing IHD, and there were certain beneficial results if the US guidelines for physical activity were satisfied.

## Introduction

Ischemic heart disease (IHD) is the first leading cause of death worldwide [1]. Every year, one-third of deaths worldwide are caused by IHD [1]. In the past, IHD was the most serious health issue and leading cause of death in older people [2]. However, this disease is not currently restricted to older people [3]. Symptoms of IHD include fast heartbeat, fainting during exercise, sweating, and fatigue; the most common are chest pain and light-headedness [2]. Engaging in physical activity is an effective and efficient strategy to prevent IHD [4]. Research shows that exercising could result in an up to three-fold IHD risk reduction [5]. The metabolic equivalent of task (MET) method is commonly used to estimate exercise intensity [6] and allows for the calculation of exercise intensity and duration.

Moreover, types of exercise are important in reducing the IHD risk [7]. Considering this, several factors have led many governments to encourage regular physical activities [8,9]. For example, the US physical activity guidelines suggest 150 min of moderate-level exercise or 75 min of weightlifting [10]. Furthermore, the US launched the ‘move your way’ campaign based on these guidelines [11]. There have been many attempts to prove the validity of the US guidelines to the public. Many studies have demonstrated that the US guidelines effectively improve public health in the general population [12–16]. However, in reality, people usually consider the type of exercise rather than its duration. This study had two aims: first, to examine the types of exercise and IHD risk. Second, to perform a meta-analysis to validate the US guidelines and determine efficient types of exercise.

## Methods

This study was conducted in accordance with the PRISMA guidelines.

### Search strategy and eligibility criteria

In this meta-analysis, six databases, including MEDLINE, EMBASE, Cochrane Library, CINAHL, Scopus, and Web of Science, were searched from January 1, 2000, to November 4, 2022. The most recent literature search was conducted on November 4, 2022, and only English-language articles were included. The Population, Intervention, Control, and Outcomes (PICO) method was used to define study characteristics.

The inclusion criteria were derived from the PICO method. Exclusion criteria were: 1) publication date before the year 2000, 2) not written in English, 3) not a primary study, 4) not a research article, and 5) not relevant to the PICO criteria. Two authors made independent extractions and discussed with the corresponding author if differences were observed.

**P:** General population

**I:** Participants who satisfied the US guidelines for physical activity

**C:** Participants who did not satisfy the US guidelines for physical activity

**O:** Reduction of IHD risk

### Categories of search terms

Physical activity [Mesh Terms and in title and abstract]: Exercise [Mesh], Physical Conditioning, Human [Mesh], Dancing [Mesh], Swimming [Mesh], Walking [Mesh], Running [Mesh], Bicycling [Mesh], Muscle Stretching Exercises [Mesh], Weight Lifting [Mesh]

Ischemic Heart Disease [Mesh Terms and in title and abstract]: Myocardial Ischemia [Mesh]

Incidence [Mesh Terms and in title and abstract]: Incidence [Mesh], Heart Disease Risk Factors [Mesh]

Human Subjects [Mesh Terms and in title and abstract]: NOT Models, Animal [Mesh] Disease Models, Animal [Mesh], Animals [Mesh]

English Only [Mesh Terms and in title and abstract]: English [la]

Years [Mesh Terms and in title and abstract]: 2000:2030 [dp]

Study Design [Mesh Terms and in title and abstract]: NOT Systematic Reviews as Topic [Mesh], Meta-Analysis as Topic [Mesh]

### Data extraction and quality assessment

After the first title/abstract screening was performed using Microsoft Excel, data was collected through full-length paper reviews [17]. Two authors made an independent assessment of the studies and discussed with the corresponding author if differences were observed. Quality assessment was performed using the Newcastle/Ottawa Scale (NOS), commonly utilized for non-randomized controlled trials [18]. Two authors made an independent assessment of the studies extracted and discussed with the corresponding author if differences were observed.

### Outcomes

We assessed exercise types and IHD risks in the primary analysis. Secondary outcomes were types and intensity of exercise that satisfied the US guidelines for physical activity and reduction in IHD risk.

### Data Analysis

We measured the results by assessing the odds ratio (OR) and 95% confidence intervals (CIs) of the studies. These standards were applied to both categorical and numerical values. A random-effect model was employed to analyze the results of a comprehensive search. The obvious and statistical analyses of the meta-analysis were estimated using the I^2^ statistics method [19]. After the data analysis process, a funnel plot was used to assess publication bias, a major bias in systematic reviews/meta-analyses [20]. Meta-analysis was performed using R Studio [21].

## Results

### Study population and selection

The literature search resulted in 7,612 articles, after eliminating duplicates. Only 270 studies satisfied the inclusion and exclusion criteria regarding their titles and abstracts. We extracted the full text of the 270 studies; however, only seven were included in the final analysis. Studies were excluded if they were based on the wrong study population (13 studies) or did not assess the types of exercise according to IHD outcomes. A total of 588,030 participants were included in the selected studies (Fig 1).

**Fig 1.**
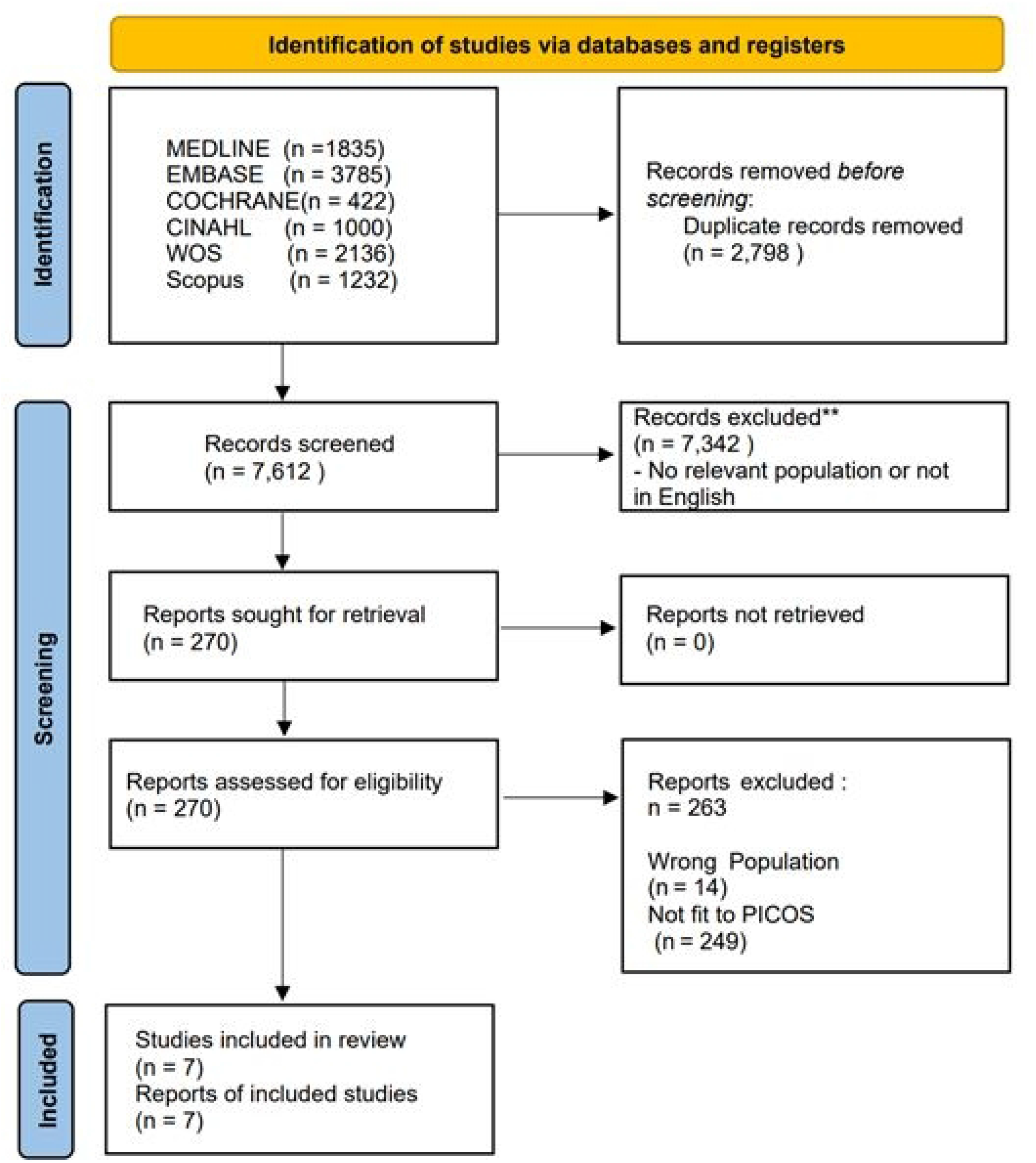
Study flow chart

### Literature review (Table 1)

**Study 1** [22]: This cohort study was conducted in 2019 in the US; the types of exercise were aerobics, weight training, and mixed exercise according to the US guidelines for physical activity. The target diseases were hypertension, myocardial infarction (MI), high cholesterol, stroke, and coronary heart disease. This study demonstrated the effect of combined moderate to vigorous physical and muscle-strengthening activity guidelines on sampled adults from the US. The expected risk of bias was recall bias because it was a self-report study.

**Table 1.** Literature review of studies.

| Study No. | Authors | Date | Study Design | Types of Exercise | Target disease | Country | Objectives |
| --- | --- | --- | --- | --- | --- | --- | --- |
| 1 | Bennie JA, De Cocker K, Teychenne MJ, Brown | 2019 | Cohort study | Aerobics, weight training, mixed | Hypertension, MI, high cholesterol, stroke, CHD | USA | To demonstrate the combined MVPA and MSA guidelines' effect on the |
|  | WJ, Biddle<br>SJH |  |  |  |  |  | sampled adults<br>from the US |
| **2** | Tanasescu<br>M,<br>Leitzmann<br>MF, Rimm<br>EB, Willett<br>WC,<br>Stampfer<br>MJ, Hu FB | 2002 | Cohort<br>study | Running,<br>jogging,<br>rowing,<br>cycling,<br>swimming,<br>racquet sports | CHD | USA | To explore the<br>type, intensity,<br>and amount of<br>exercise in<br>relation to the<br>risk of coronary<br>heart disease in<br>men |
| **3** | Hu G,<br>Tuomilehto<br>J, Borodulin<br>K, Jousilahti<br>P | 2020 | Cohort<br>study | Occupational<br>exercise,<br>walking or<br>cycling to and<br>from work,<br>LTPA | CHD | Finland | To prove<br>associations<br>between types<br>of physical<br>activity and a<br>decade risk of<br>CHD using the<br>FRS |
| **4** | Lovasi GS,<br>Lemaitre<br>RN,<br>Siscovick | 2007 | Case-control<br>study | Walking,<br>gardening,<br>exercise<br>cycling, | MI | USA | To investigate<br>the association<br>between the<br>amount of |
|  | DS, Dublin<br>S, Bis JC,<br>Lumley T, et<br>al. |  |  | bowling,<br>calisthenics,<br>exercise<br>machines,<br>dancing,<br>golfing, weight<br>training,<br>swimming,<br>aerobics,<br>hiking |  |  | LTPA and<br>reduction of MI<br>risk |
| 5 | Chomistek<br>AK,<br>Henschel B,<br>Eliassen AH,<br>Mukamal<br>KJ, Rimm<br>EB | 2016 | Cohort<br>study | Jogging,<br>running,<br>bicycling,<br>swimming,<br>tennis | CHD | USA | To assess the<br>association<br>between the<br>volume of<br>LTPA measured<br>by MET-hours/wk |
| 6 | Porter AK,<br>Schilsky S,<br>Evenson<br>KR, Florido<br>R, Palta P, | 2019 | Cohort<br>study | Aerobics,<br>basketball,<br>bicycling,<br>calisthenics,<br>golf walking, | Cardiovascular<br>disease | USA | To examine the<br>relationships<br>between<br>participation in<br>the types of |
|  | Holliday<br>KM, et al. |  |  | racquet sports,<br>running,<br>swimming,<br>walking,<br>weight training |  |  | exercise and<br>incident<br>myocardial<br>ischemia |
| 7 | Fransson E,<br>De Faire U,<br>Ahlbom A,<br>Reuterwall<br>C, Hallqvist<br>J, Alfredsson<br>L | 2004 | Cohort<br>study | Repetitive<br>lifting at work,<br>heavy lifting at<br>work,<br>household<br>work | MI | Sweden | For the<br>estimation of<br>household<br>work, exercise,<br>and<br>occupational<br>physical<br>activity, and<br>risk of acute MI<br>relatively. The<br>potential<br>interaction of<br>physical activity<br>was examined |

Abbreviations: CHD, coronary heart disease; FRS, Framingham risk score; LTPA, Leisure- Time Physical Activity; MET, metabolic equivalent of task; MI, myocardial infarction; MVPA moderate to vigorous physical activity; MSA, and muscle strengthening activity

**Study 2** [23]: This cohort study was conducted in 2002 in the US. The types of exercises were running, jogging, rowing, cycling, swimming, and racquet sports. The target disease was coronary heart disease. The study aimed to explore the type, intensity, and amount of exercise with respect to the risk of coronary heart disease in men. The expected risk of bias was misclassification bias because it was a self-report study.

**Study 3** [24]: This cohort study was conducted in 2020 in Finland. The types of exercises were occupational exercise, walking or cycling to and from work, and Leisure-Time Physical Activity (LTPA). The target disease was coronary heart disease. The study aimed to prove the association between types of physical activity and a decade risk of MI using the Framingham risk score. The expected risk of bias was recall bias because it was a self-report study.

**Study 4** [25]: This cohort study was conducted in 2007 in the US, and the types of exercise were walking, gardening, exercise cycling, bowling, calisthenics, exercise machines, dancing, golfing, weight training, swimming, aerobics, and hiking. The target disease was MI. The study investigated the association between the amount of LTPA and reduction of MI risk. The expected risk of bias was recall bias because it was a self-report study.

**Study 5** [26]: This cohort study was conducted in 2016 in the US. The types of exercise considered were walking, gardening, cycling, bowling, calisthenics, exercise machines, dancing, golfing, weight training, swimming, aerobics, and hiking. The target disease was MI. The study aimed to investigate the association between the amount of LTPA and MI risk reduction. The expected risk of bias was recall bias because it was a self-report study. The authors also reported a possible underestimation of the association with MI risk for survivors.

**Study 6** [27]: This cohort study was conducted in 2019 in the US. The exercises considered were aerobics, basketball, bicycling, calisthenics, golf walking, racquet sports, running, swimming, walking, and weight training. The target disease was MI. The study assessed the association between the volume of LTPA measured by the MET-hours/week. The expected risk of bias was recall bias because it was a self-report study.

**Study 7** [28]: This cohort study was conducted in 2004 in Sweden. The types of exercise were repetitive lifting at work, heavy lifting at work, and household work. The target disease was MI. The study examined the relationship between participating in the types of exercise and incidents of myocardial ischemia. The expected risk of bias was reporting and misclassification bias because it was a self-report study.

Abbreviations: CHD, coronary heart disease; FRS, Framingham risk score; LTPA, Leisure-Time Physical Activity; MET, metabolic equivalent of task; MI, myocardial infarction; MVPA moderate to vigorous physical activity; MSA, and muscle strengthening activity

## Quality assessment

The NOS was utilized to assess the quality of the selected studies.

### Study outcomes

Concerning the types of exercise that satisfied the US guidelines for physical activity, there were 11 values for aerobics, 6 for lifting, and 4 for mixed exercises. Regarding the details of physical activity, there were 4 values for aerobics, 1 for dancing, 1 for hiking, 2 for swimming, 1 for basketball, 1 for running, 1 for calisthenics, 1 for exercise machines, 3 for weight training, 1 for biking, 1 for bowling, and 1 for mixed exercises.

The study design analysis included 9 values for case-control studies and 12 for cohort studies. Target diseases comprised 12 values for MI; 4 for IHD, heart failure, or stroke; 3 for myocardial ischemia; and 2 for stroke. Trim and Fill analysis and a funnel plot analysis were performed to assess publication bias; however, none was observed (S1 Table 1).

### Primary outcomes

Regarding heterogeneity of the studies, 96.1% of the included studies had a very high level of heterogeneity when the random effects model was used. The overall effect of types of physical activity on IHD was OR: 0.744 (95% CI: 0.740–0.748). The ORs for the standards used were 0.813 (95% CI: 0.779–0.849) for aerobics, 0.917 (95% CI: 0.823–1.023) for weight training, and 0.681 (95% CI: 0.637–0.727) for mixed exercises. For the target diseases, the OR for hypertension was 0.752 (95% CI: 0.647–0.875), 0.819 (95% CI: 0.716-0.938) for high cholesterol, 0.703 (95% CI: 0.634–0.779) for MI, 0.788 (95% CI: 0.751– 0.826) for IHD, 0.788 (95% CI: 0.715–0.869) for stroke, and 0.846 (95% CI: 0.788–0.909) for heart failure. Across countries, the OR for studies in the US was 0.773 (95% CI: 0.742– 0.805), 0.702 (95% CI: 0.619–0.797) for those in Finland, and 0.779 (95% CI: 0.678-0.896) for those in Sweden (Fig 2). The funnel plot and trim and fill analysis showed no publication bias (S1 Fig).

**Fig 2.**
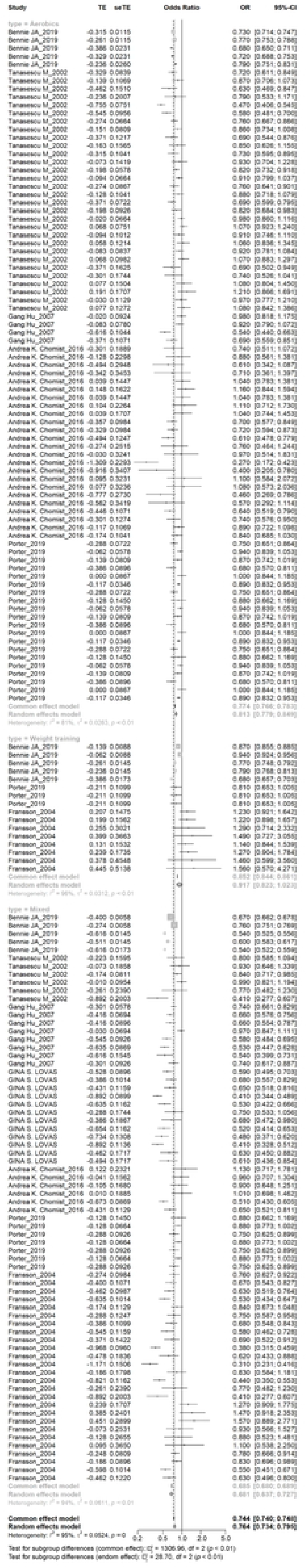
Forest plot generated by types of exercise. TE, estimated treatment effect; seTE, standard error of treatment estimate; OR, odds ratio; CI, confidence interval

### Secondary outcomes

Regarding heterogeneity of the studies, 16.1% of the studies showed heterogeneity. The ORs of aerobics, weight training, and mixed exercises were 0.607 (95% CI: 0.469–0.787), 0.569 (95% CI: 0.388–0.833), and 0.585 (95% CI: 0.370–0.924), respectively (Fig 3).

**Fig 3.**
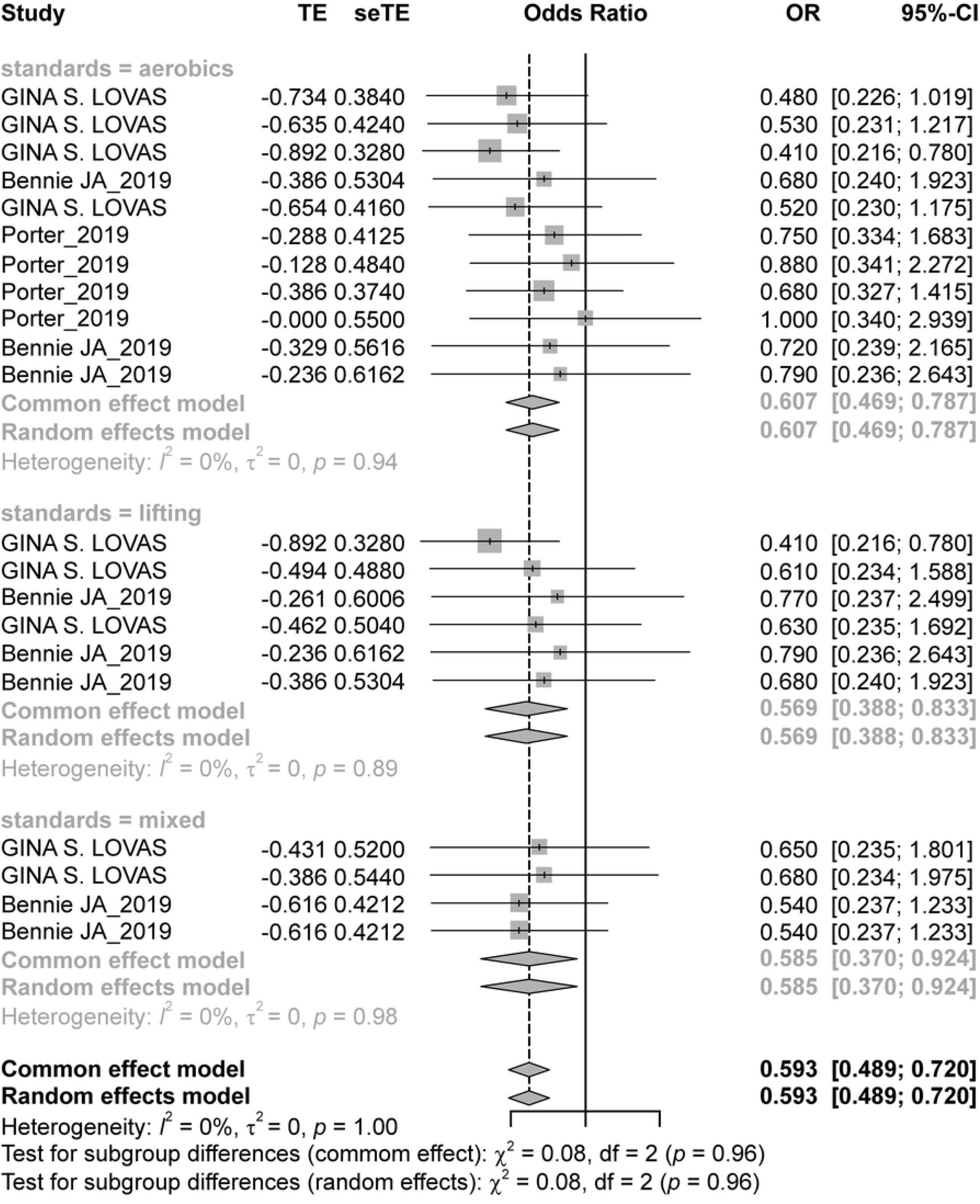
Forest plot generated by standard satisfaction. TE, estimated treatment effect; seTE, standard error of treatment estimate; OR, odds ratio; CI, confidence interval

For target diseases, the ORs for IHD were 0.515 (95% CI: 0.401–0.662), 0.777 (95% CI: 0.540–1.116), and 0.630 (95% CI: 0.356–1.114), respectively (Fig 4).

**Fig 4.**
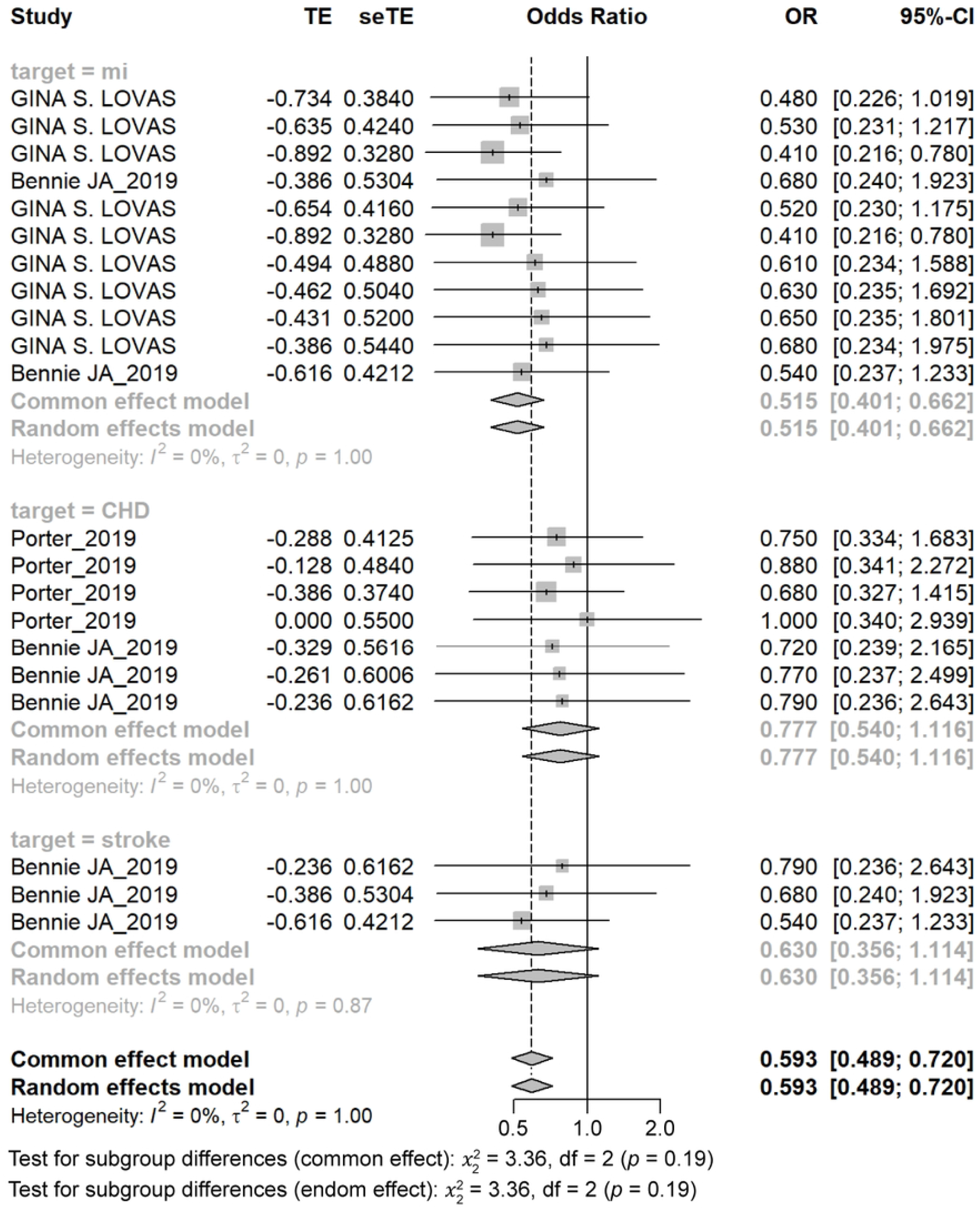
Forest plot generated by target disease from studies that satisfied the US guidelines. MI, myocardial infarction; CHD, coronary heart disease; TE, estimated treatment effect; seTE, standard error of treatment estimate; OR, odds ratio; CI, confidence interval

For study designs, the OR for case-control studies was 0.503 (95% CI: 0.383–0.661) and 0.703 (95% CI: 0.533–0.927) for cohort studies. When exercises were included in the model, the OR for LTPA was 0.503 (95% CI: 0.383–0.661); 0.733 (95% CI: 0.444–1.212) for aerobics; 0.880 (95% CI: 0.341–2.272) for basketball; 0.680 (95% CI: 0.327–1.415) for running; 1.000 (95% CI: 0.340–2.939) for swimming; 0.738 (95% CI: 0.384–1.421) for weight training; and 0.540 (95% CI: 0.301–0.968) for mixed exercises.

## Discussion

Our study showed that aerobics and weight training have beneficial effects on IHD. Without the US guidelines for physical activity, there was a high level of heterogeneity in the effects of physical activity by type, with overall effects of 0.764 (OR; 95% CI: 0.737–0.795). No publication bias was observed. However, after applying the US guidelines for physical activity, there was a low level of heterogeneity in its effects and an observed OR of 0.515 (95% CI: 0.401–0.662) for MI. A significant reduction in the heterogeneity of the studies after applying the US guidelines suggests that these recommend certain levels of exercise intensity and include the duration of exercise irrespective of the types of exercise. There was no significant difference among countries. In a previous related study, increased physical activity was found to be associated with lower blood pressure in hypertensive individuals, increased high-density lipoprotein cholesterol levels in a dose-response manner, and reduced incidence of diabetes [29]. The health benefits of physical activity can be achieved by engaging in moderate-intensity physical activity (brisk walking) for at least 30 min per day, 5 days per week, or vigorous activity (jogging) for 20 min or more, 3 days per week. Combinations of the two types of activity can also be performed. Furthermore, a mendelian randomization study reported that genetically predicted self-reported vigorous physical activity was significantly associated with a lower risk of MI (OR: 0.24, 95% CI: 0.08–0.68; p-value: 0.007) [30]. Additionally, those results were consistent after the sensitivity analysis. The INTERHEART study demonstrated that regular exercise reduced the risk of MI (OR: 0.86) [31]. A different study, which followed 84,129 women who engaged in moderate or vigorous exercise for over 30 min per day, reported a relative risk of 0.17 [32].

The strength of the current study was that the beneficial effects of physical activity on cardiovascular health were reported with the US guidelines for physical activity. The level, types, and intensity of physical activity were associated with the regular and designed guidelines for physical activity.

This study had some limitations. First, there was a possibility of length bias in the mixed session; therefore, the OR may have been lower. Second, we classified the exercise types, but did not account for the baseline and post-exercise development of muscle or body fat. For example, a research article published in 2015 found that quadriceps strength was highly correlated with reduced mortality due to coronary artery disease [33].

## Conclusions

Although many research papers have focused on exercise and its efficiency in MI risk reduction, the association between this reduction and exercise type has not been well-researched. This study conducted a meta-analysis to assess which types of exercise efficiently reduce MI and assess efficiency according to the US guidelines for physical activity. This study concluded that beneficial results were obtained if the US guidelines for physical activity were followed. This also shows the need for the implementation of health policies regarding regular physical activity, which could aid in preventing an increase in IHD incidence and its negative outcomes, especially MI. Community-based programs for regular physical activity are reportedly effective in reducing the IHD risk [11]. Therefore, community and national programs, such as the “move your way” program, should be implemented to improve the population’s cardiovascular health [11].

Further research should aim to show the efficiency of IHD risk reduction, as previous case-control studies with experimental design have shown the likelihood of patients with IHD benefitting from regular exercise practice.

## Data Availability

The data underlying the results presented in the study are available from (include the name of the third party and contact information or URL).

## Author contributions

**Jaewoo Cha**: the conception and design of the study, or acquisition of data, or analysis and interpretation of data, drafting the article or revising it critically for important intellectual content.. **Kwan Hong:** final approval of the version to be submitted., **Jeehyun Kim:** final approval of the version to be submitted.

## Supporting information

**S1 Table. Newcastle-Ottawa Scale of study assessment**.

**S1 Fig. Funnel plot of study**

## Notes

### Competing Interest Statement

The authors have declared no competing interest.

### Funding Statement

The funders had no role in study design, data collection and analysis, decision to publish, or preparation of the manuscript.

